# SARS-CoV-2 Omicron BA.5: Evolving tropism and evasion of potent humoral responses and resistance to clinical immunotherapeutics relative to viral variants of concern

**DOI:** 10.1101/2022.07.07.22277128

**Authors:** Anupriya Aggarwal, Anouschka Akerman, Vanessa Milogiannakis, Mariana Ruiz Silva, Gregory Walker, Andrea Kidinger, Thomas Angelovich, Emily Waring, Supavadee Amatayakul-Chantler, Nathan Roth, Germano Coppola, Malinna Yeang, Tyra Jean, Charles Foster, Alexandra Carey Hoppe, C. Mee Ling Munier, David Darley, Melissa Churchill, Damian Starck, Daniel Christ, Gail Matthews, William Rawlinson, Anthony D. Kelleher, Stuart Turville

## Abstract

Genetically distinct viral variants of severe acute respiratory syndrome coronavirus 2 (SARS-CoV-2) have been recorded since January 2020. Over this time global vaccine programs have been introduced, contributing to lower COVID-19 hospitalisation and mortality rates, particularly in developed countries. In late 2021, the Omicron BA.1 variant emerged, with substantially different genetic differences and clinical effects from other variants of concern (VOC). This variant demonstrated higher numbers of polymorphisms in the gene encoding the Spike (S) protein, and it has displaced the previously dominant Delta variant. Shortly after dominating global spread in early 2022, BA.1 was supplanted by the genetically distinct Omicron lineage BA.2. A sub-lineage of BA.2, designated BA.5 has now started to dominate globally, with the potential to supplant BA.2. To address the relative threat of BA.5, we determined infectivity to particle ratios in primary nasopharyngeal samples and expanded low passage isolates in a well characterised, genetically engineered ACE2/TMPRSS2 cell line. We then assessed the impact of BA.5 infection on humoral neutralisation *in vitro*, in vaccinated and convalescent cohorts, using concentrated human IgG pooled from thousands of plasma donors, and licensed monoclonal antibody therapies. The infectivity of virus in primary swabs and expanded isolates revealed that whilst BA.1 and BA.2 are attenuated through ACE2/TMPRSS2, BA.5 infectivity is equivalent to that of an early 2020 circulating clade and has greater sensitivity to the TMPRSS2 inhibitor Nafamostat. As with BA.1, we observed BA.5 to significantly reduce neutralisation titres across all donors. Concentrated pooled human IgG from convalescent and vaccinated donors had greater breadth of neutralisation, although the potency was still reduced 7-fold with BA.5. Of all therapeutic antibodies tested, we observed a 14.3-fold reduction using Evusheld and 16.8-fold reduction using Sotrovimab when neutralising a Clade A versus BA.5 isolate. These results have implications for ongoing tracking and management of Omicron waves globally.

## Introduction

At the beginning of November 2021, the VOC Omicron BA.1 surged globally with close to 4 million infections per day reported by mid-January. This variant was then supplanted by the genetically divergent BA.2 variant, which represented over 80% of cases reported worldwide by mid-April of 2022. In June 2022, three lineages derived from BA.2 were starting to dominate (. These included BA.2.12.1, BA.4 and BA.5. BA.4 and BA.5 share amino acid substitutions (compared to BA.2) L452R, F486V, and R493Q in the Spike receptor binding domain (RBD) whereas BA.2.12.1 is the only variant with the L452Q change. Across several areas globally, BA.5 is spreading preferentially over BA.2.12.1 and BA.4. The determinants of preferential spread are complex and must take into account many variables, including the prevalence of infection and/or vaccine coverage and the time from that latter antigenic exposure. In addition to the population level of immunity, the mechanism of viral entry and changes thereof may significantly influence viral tropism and subsequent disease severity even within previously vaccinated populations. For instance, the Delta variant had significant tropism for the ACE2-TMPRSS2 pathway and this pathway is associated with infection of the lung and disease severity in animal models ^1-6^. In contrast, Omicron BA.1 diverged from this pathway with a tropism trajectory towards the upper respiratory tract ^3^. The mechanism for Omicron favouring the upper respiratory tract is presently hypothesised to be the switch from TMPRSS2 to another serine or cysteine protease either present at the plasma membrane or enriched within the endolysosomal compartment ^3,7-9^. Whilst BA.2 has similar tropism to BA.1, recent studies on BA.5 and related lineages bearing L452 polymorphisms highlight a shift in tropism back to pre-Omicron lineages, with potential increase in disease severity and infection within lung tissue observed in animal models (**doi:** https://doi.org/10.1101/2022.05.26.493539).

Recently we developed a rapid and sensitive platform for the isolation and characterisation of SARS-CoV-2 variants with respect to their relative transmission threat in previously infected and vaccinated populations ^7^. This platform rapidly feeds back three key observations with respect to early characterisation of viral variants in primary nasopharyngeal samples. Firstly, it enables neutralisation studies on primary clinical viral isolates. Secondly, it determines which immunotherapeutics retain potency. Finally, it can resolve subtle changes in tropism towards or away from from the ACE2-TMPRSS2 pathway of primary clinical isolates by the increase or decrease of the viral infectivity to particle ratio. In the latter setting, this system could not only map increase usage of TMPRSS2 by Delta in primary nasopharyngeal swabs, but also readily demonstrate the decreased use of TMPRSS2 by Omicron BA.1. As this can be done with diagnostic primary samples, it can reveal tropism changes when a variant starts expanding within a community in real time.

Through using individual serum samples from 74 patients recruited to ADAPT^10^, a community-based cohort of approximately 200 patients followed from the time of diagnosis during all waves of infection in Australia, we tested a continuum of responses ranging from triple vaccinated donors, convalescent donors post vaccination and Omicron breakthrough infections of triple vaccinated individuals. To assess breadth across variants, we tested live virus neutralisation potency against the pre-Omicron clades A2.2, Beta and Delta alongside the Omicron lineages BA.1, BA.2 and BA.5. We then tested 13 polyclonal human hyperimmune IgG batches that constitute pools of thousands of primarily US plasma donors collected in late 2021 prior to the onset of the global Omicron BA.1 wave (Figure 1E). This latter analysis establishes the extent of immune evasion at the population level at that time period, as the IgG is comprised of all plasma donors irrespective if they are convalescent and/or vaccinated. Alongside patient sera, we also tested clinical grade Xevudy (sotrovimab) and Evusheld (tixagevimab and cilgavimab) for changes in potency across all above listed variants. Finally, with overlapping waves of BA.2 and BA.5 infection within Australia at the time of writing, we then determined the infectivity to particle ratios of virus within primary nasopharyngeal swabs and furthermore established the mode of entry of BA5 versus other Omicron and pre-Omicron lineages.

**Figure 1.**
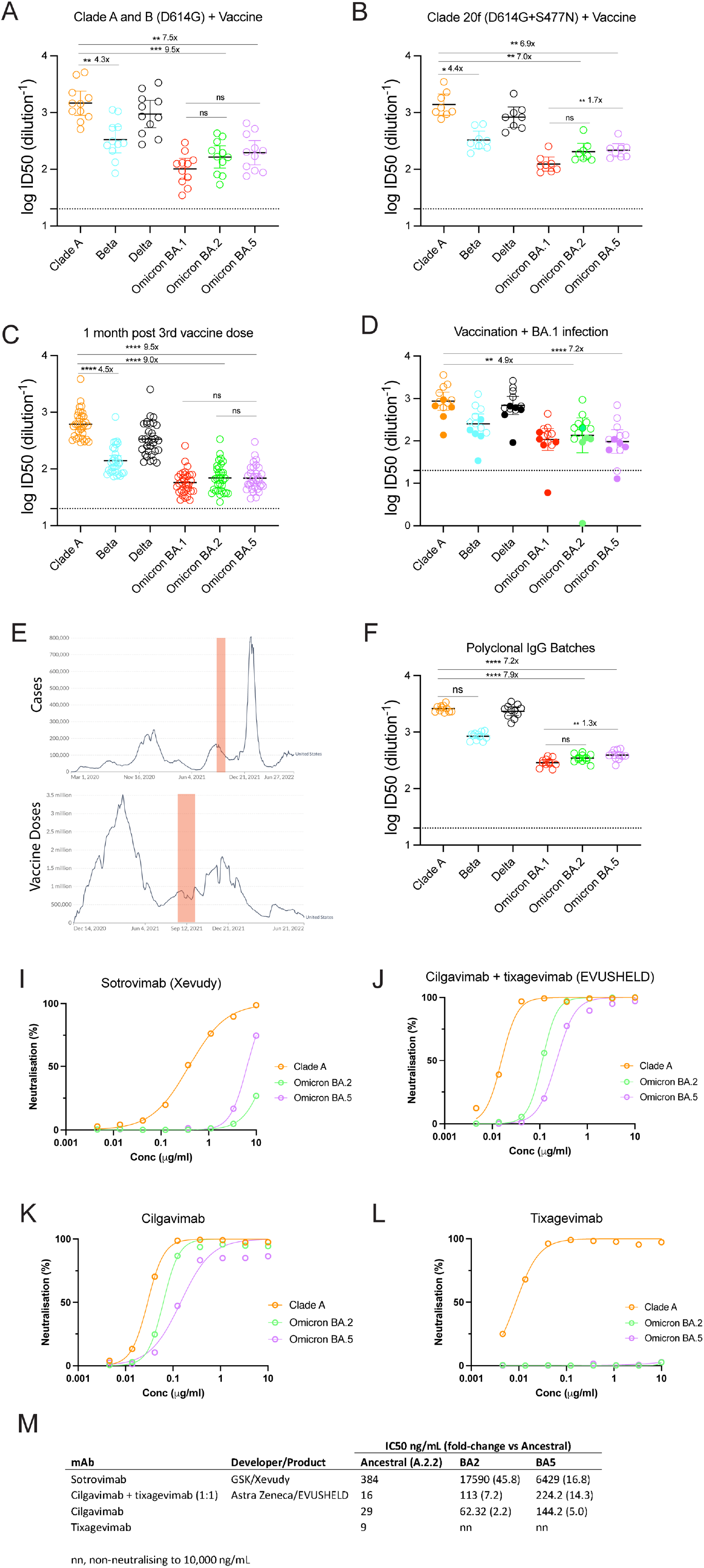
Humoral neutralisation of clinical SARS-CoV-2 variants in convalescent and vaccinated donors, and pooled concentrated human IgG plasma samples. Neutralisation assays were performed in a high-throughput format in HAT-24 cells using live virus isolates from the variants of concern Delta (B.1.617.2), Beta (B.1.351), Omicron BA.1, Omicron BA.2 and Omicron BA.5 and the ancestral Wuhan-like virus with the original D614 background (A2.2) as a control. ID50 neutralisation titres presented for 6 live variants for vaccinated donors from **(A)** Clade A and B (D614G) (first wave), **(B)** Clade 20F (D614G + S477N) (the second wave), **(C)** healthy donors one month after third dose of vaccination, **(D)** Vaccinated (open circles) or unvaccinated donors (solid circles) infected with Omicron BA.1, **(E)** SARS-CoV-2 cases (upper panel) and vaccination doses (lower panel) across the United States population where US plasma donors were sourced for polyclonal immunoglobulins (Poly-Ig). Shaded areas highlight the time at which the plasma donors units were used for the generation of Poly-Ig (greater than 10,000 donors were pooled for each batch tested) and **(F)** concentrated polyclonal IgG from either convalescent and vaccinated donors. Data in **(A-D and F)** indicates the mean ID50 of technical replicates for individual samples (circles) with the geometric mean and 95% confidence interval shown for each variant. Dotted line represents limit of detection (LOD). Fold change reductions in ID50 neutralisation titres compare variants of concern to the ancestral variant, as well as to Omicron BA.1 where indicated. *p<0.05, **p<0.01, ***p<0.001, ****p<0.0001 for Kruskal Wallis test with Dunn’s multiple comparison test. **I-F**. Neutralization activity of monoclonal antibodies Sotrovimab **(I)**, Cilgavimab and Tixagevimab cocktail **(J)**, Cilgavimab alone **(K)** and Tixagevimab alone **(L)** against ancestral A2.2, Omicron BA.2 and Omicron BA.5 **(M**) IC50 values (ng/ul) and fold change relative to ancestral A2.2 for the monoclonal antibodies against Omicron BA.2 and BA.5. Antibodies used herein were clinical grade batches.

## Results and discussion

### Humoral evasion for Omicron lineages BA.1, BA.2 and BA.5 relative to early clade A, Beta and Delta variants

The ability of Omicron lineages and other variants (Beta and Delta) to evade neutralising antibody responses was assessed using our rapid 20-hour live virus neutralisation platform (R-20) ^7^. Initially we examined neutralisation responses in sera from convalescent donors from early clade infections who had subsequently been vaccinated with either the BNT162b or ChAdOx1 vaccine (Figure 1A and B and Supplementary Table 1; presented together as a single “vaccinated” group). While the infected and vaccinated individuals are observed to have potent neutralisation titres to the ancestral strain (clade A), we observed a 7 to 15-fold reduction in neutralisation against all Omicron sub-lineages (15.6-fold for BA.1, p<0.0001; 9.5-fold for BA.2, p<0.001; 7.5-fold for BA.5, p<0.01) compared to a 1.5 and 4.3-fold decrease observed for Delta and Beta relative to Clade A, respectively (p>0.05). Similar reductions in neutralisation were also seen in sera from individuals infected during the second wave (Fig 1B) or in those who were SARS-CoV-2 naïve but triple vaccinated (Figure 1C; Supplementary Tables 1&2). Whilst we had previously observed greater fold reductions (approximately 2-fold higher) using the same sera to Omicron BA.1 ^7^, changes in viral expansion would account for this observation, as the cells used enabled harvesting of virus within a 24-hour window and therefore limited the accumulation of non-viable viral particles (we have calculated the half-life of SARS-CoV-2 at room temperature to be 1.4 days).

Similar results were obtained when we tested neutralisation responses against thirteen polyclonal human IgG batches comprised of more than ten thousand pooled plasma donors per batch collected during late 2021, following the peak of the Delta wave but just prior to the BA.1 Omicron wave (Figure 1F; Supplementary Table 1). There was a 7 to 9-fold reduction in neutralisation against Omicron lineages (9.4-fold for BA.1, p<0.0001; 7.9-fold for BA.2, p<0.0001 and 7.2-fold for BA.5, p<0.001) compared with 3.2-fold (p>0.05) and 1.1-fold decrease (p>0.05) for Beta and Delta, respectively. Across most patient samples, we observed no significant differences when comparing the Omicron lineages BA.1, BA.2 and BA.5.

Whilst the above samples give a population snapshot of antibody potency and breadth from donors in 2021, we then turned to serum samples collected from the ADAPT cohort where donors were previously triple vaccinated but were then infected during the BA.1 wave. Given the large proportion of the vaccinated population were those infected globally with Omicron BA.1, our aim was to determine the potential increase in breadth towards Omicron lineages following BA.1 infection. Across all donors, we observed greater neutralisation potency to pre-Omicron viral variants, with neutralisation titres to Omicron lineages similar to that observed with SARS-CoV-2 naïve triple vaccinated donors (Figure 1D; Supplementary Table 1). Two donors did demonstrate significant neutralisation titres to all Omicron lineages, but they were lower than their observed responses to pre-Omicron variants.

### Evusheld and Sotrovimab activity against Omicron clinical BA.2 versus BA.5 isolates

The currently clinically utilised monoclonal antibodies (mAbs), Evusheld and Sotrovimab, were assessed for neutralisation capacity primarily against the Omicron variants BA.2 and BA.5, as they currently represent the dominant variants within the community. These therapies may need to be used in individuals that have not mounted a vaccine response from therapeutic induced or pre-existing immunodeficiencies. Whilst activity was retained using Evusheld for BA.2, we did observe a drop in potency when testing BA.5. Curiously, whilst there was no detectable activity of Sotrovimab against BA.2, we did observe activity towards BA.5 but at lower potency compared to the ancestral clade A2.2 variant (Figure 1IM).

All Omicron variants contain significant substitutions within their RBD site relative to the ancestral variant. Therefore, fluctuations in potency in antibodies against the RBD (e.g. Evusheld) are to be expected. Furthermore, the Evusheld binding sites cover the unique RBD polymorphisms (L452R, F486V, and R493Q) that distinguish the BA.5 spike glycoprotein from the BA.2 spike. In contrast, the class 3 antibody Sotrovimab targets a highly conserved region of the sarbecovirus RBD ^11^ and based on its neutralising epitope should retain activity against both BA.2 and BA.5. With no detectable activity towards BA.2 even at the highest dose tested and now detectable activity towards BA.5, the above RBD changes outside of the Sotrovimab epitope support a conformation change in the Spike glycoprotein that now renders BA.5 susceptible to neutralisation. Given the continual emergence of new variants on a periodic basis and the continued high prevalence of infection globally, the need for therapeutic treatments still remains high and is of high priority for those who are immunocompromised. Whilst the retention of neutralising activity by Sotrovimab and Evusheld against Omicron BA.5 is promising, the continued trajectory of lower monoclonal antibody potency remains a concern, and the development of new and improved monoclonal antibody modalities and alternative therapies is still urgently warranted. As with treatment of other RNA viruses (e.g. HIV-1 and HCV), combination therapies are likely to limit the appearance of therapeutic resistant variants that can arise within the clinic.

### Shifting tropism of the BA.5 Omicron variant

In the pre-Omicron era, we developed a hyper-permissive ACE2-TMPRSS2 Hek293T cell line (HAT-24) with culture sensitivity for detection approaching that of diagnostic PCR ^7,10^. During the Delta wave, virus could be cultured 75% of the time from clinical swabs with diagnostic PCR Ct values above 30 ^7^. In using this cell type, we curated clinical samples that were PCR positive and cryopreserved them at -80°C within 24 hours. Using this material, we established linear regressions of infectivity (TCID50/ml) versus particle number (viral load-diagnostic PCR Ct value) across a number of clinical specimens with increasing levels of viral load. Using this approach, we could readily resolve the increased infectivity to particle ratio of an early circulating variant against that of a variant with increased tropism towards the ACE2-TMPRSS2 pathway, (See Delta Fig 2A and 2E). Although, with the emergence of Omicron BA.1, both ourselves and others have observed that the Omicron lineage BA.1 uses the ACE2-TMPRSS2 pathway poorly. This is consistent with our viral outgrowth results using this ACE2-TMPRS2 cell line (Fig 2A and G). Whilst transmissibility of pre-Omicron variants was linked closely to TMPRSS2 use, the increased transmissibility of Omicron lineages BA.1 and BA.2 support the use on an alternate protease either at the membrane, like TMPRSS2 or via the endosome through cysteine proteases^3,7-9^. Following the BA.2 wave in Australia, we are now experiencing increasing cases of BA.5. Using the above cell line, we collected samples over a time period when both BA.2 and BA.5 were in high prevalence within the community. In this setting, any immunity in the community that could influence the results was controlled for by sampling within the same time window. In testing collections of BA.2 and BA.5 samples, it was immediately apparent that by culture of the virus swab alone, BA.2 could be readily differentiated from BA.5. As was observed with BA.1, BA.2 was poorly infectious in the HAT-24 ACE2-TMPRSS2 cell line. This can be easily visualised after three days of culture with limited cytopathic effects accumulating within this cell line (Figure 2C), even when samples with high viral loads were used. In contrast, BA.5 could mediate not only extensive cytopathic effects (Fig 2D) but was also significantly more infectious per diagnostic Ct value compared to the co-circulating BA.2 parental variant (Fig 2E and F). In comparison with prior circulating variants the y intercept of the linear regression of Ct v infectivity for BA.5 sits below that of Delta but alongside that of an early circulating variant in 2020 (Fig 2E). However, for the latter comparisons, we need to take into account that these isolates were tested when the community in Australia was primarily unvaccinated. To further interrogate the increase of infectivity per Ct value, we expanded pre-Omicron and Omicron variants at the same time using the same VeroE6-TMPRSS2 cell line. The latter cell line overexpresses TMPRSS2, so even variants that poorly utilise TMPRSS2 can be expanded under similar conditions over 24 hours. With respect to RNA copies per ml, this cell line could generate viral stocks with similar particle numbers across all variants (Supplementary table S3). The only exception was Delta, which in this line exhibited greater levels of cytopathic effect (Fig 2D) and as such particle numbers were lower (Supplementary table S3). When testing these samples within the HAT-24 cell line and accounting for particle input, we could readily recapitulate the infectivity to particle ratio we had previously established using primary clinical swabs (Fig 2F).

**Figure 2.**
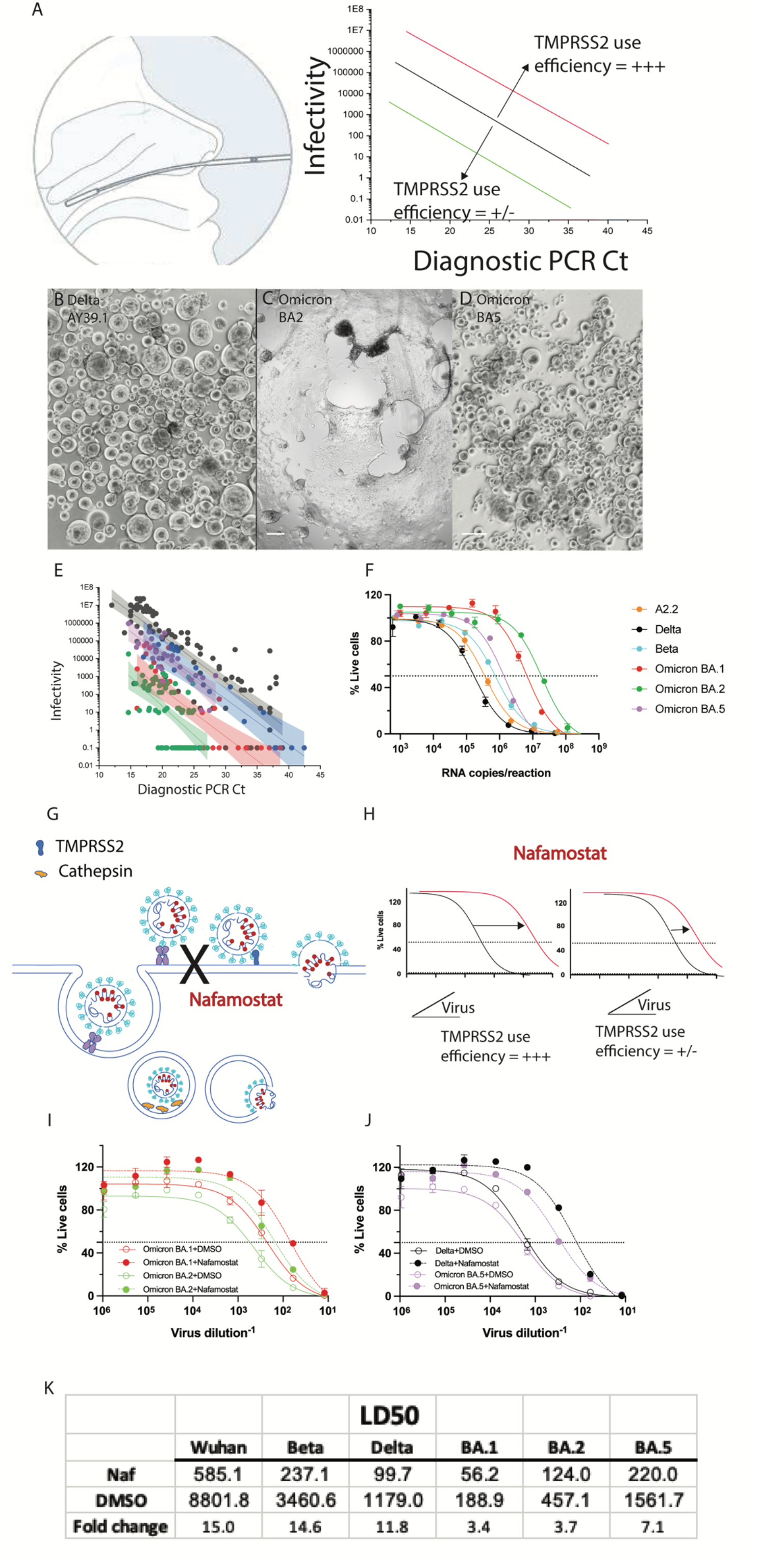
Changing tropism of the BA.5 variant relative to BA.1, BA.2 and pre-Omicron variants. **(A)** Shift in linear regression between virus infectivity and diagnostic PCR Ct values gives a measure of TMPRSS2 use by individual variants **(B-D)** Primary nasopharyngeal swabs were used to inoculate the HAT-24 cell line. All swabs are representative of high viral loads range from diagnostic PCR Ct values of 17 to 19. Cultures of **(B)** AY39.1 (one of the last detected Delta lineages), **(C)** BA.2 and **(D)** BA.5 imaged 72 hours post-infection. Of note, only BA.2 and BA.5 were from samples collected at the same time period. Scale bars represent 100 μm. **(D)** Infectivity (TCID50/ml) is presented against original diagnostic PCR Ct values for BA.2 and BA.5. Shading represents the 95% confidence intervals for each linear regression. As a comparison, earlier tested samples from Delta and an early 2020 circulating clade are also presented. Of note, only BA.2 and BA.5. samples have been collected at the same time period. **(F)** Three early clade variants (A.2.2, Beta and Delta) and three Omicron sub-lineages were grown within a 24-hour time frame under identical culture conditions (i.e. expanded at the same time, with the same MOI and harvest time). Virion particle counts were then determined using quantitative RT-PCR. Titres were then determined overnight using the R-20 assay to establish infectivity per variant per viral particle. In brief, the HAT-24 cell line rapidly develops cytopathic effects overnight, in a dose-dependent manner that can be enumerated by nuclei counts using high content microscopy. Infectivity can then be determined by calculating the 50% death of cells within the culture (LD50). Each point and error bar represent the mean and standard deviation respectively of four technical replicates. (G) Schematic showing the effect of TMPRSS2 inhibitor Nafamostat on virus entry at the cell membrane. (H) Efficiency of TMPRSS2 usage by the virus can be determined by observing a shift in LD50 when virus is titered in the presence of saturating levels of Nafamostat. Virus titrations of **(I)** Delta and BA.5 and **(J)** BA.1 and BA.2 were carried out in the presence of 20 μM Nafamostat (dashed line) or DMSO (solid line) and (**K**) change in LD50 values was used as a measure of TMPRSS2 sensitivity. Data is representative of two independent experiments. Images in (**B-D**) are representative of >20 primary samples with high viral loads. Data in (**F**) is representative of three independent low passage expansions of the 6 isolates used.

To understand the mechanism whereby the HAT-24 cell line can readily rank primary isolates based on increasing efficiency of TMPRSS2 use, we determined the expression of TMPRSS2 on this cell line versus that of the SARS-CoV-2 permissive cell line Calu3. In this setting, we observed that the HAT-24 cell line abundantly expresses ACE2, but expresses low levels of TMPRSS2. In contrast, Calu3 cells were ACE2 low but TMPRSS2 high (Supplementary Fig 1). Therefore, we conclude, the rate limiting entry step for the HAT-24 line is TMPRSS2 and when variants poorly utilise this serine protease, it can be readily resolved in culture even with primary swab samples.

To further investigate the relative usage of TMPRSS2 by Omicron lineages, we performed titrations of virus stocks in the presence of saturating amounts of Nafamostat (20 μM), a well characterised TMPRSS2 inhibitor (Fig 2G&H). Delta was used as control in this setting as this variant has previously been shown to use TMPRSS2 efficiently ^3,12^. The impact of Nafamostat on infectivity was then examined by determining the virus dilution that induces 50% cell death in culture (LD50) in the presence of this inhibitor (Fig 2H, I&J). As observed with Delta and Omicron BA1 and BA2, the LD50 is influenced by the efficiency of TMPRSS2 usage by each isolate. However, it must be noted that all SARS-CoV2 isolates can enter cells through endocytosis and as such infectivity still persists in the absence of TMPRSS2. To confirm a proportion of the LD50 values of each isolate were a function TMPRSS2 use, we used saturating levels of Nafamostat (determined through initial titrations using Delta). In this setting the greater the drop of the LD50 in the presence of the TMPRSS2 inhibitor Nafamostat, the more the isolate was using TMPRSS2 in relation to viral entry. Using this approach, we observed an an 11-fold drop in infectivity for Delta versus a 3-fold drop for both BA.1 and BA.2 in the presence of Nafamostat (Fig 2K). This is consistent with Delta’s efficient use of TMPRSS2 for viral entry compared to the poor TMPRSS2 use by Omicron lineages BA1 and BA2. For BA5, we observed a 7.0-fold drop in infectivity of BA.5 in presence of Nafamostat, which confirms the increased infectivity to particle ratio over other Omicron lineages in the HAT-24 line to be primarily related to more efficient TMPRSS2 use. Importantly, extensive observations by independent teams for pre-Omicron and Omicron lineages BA.1 and BA.2 readily support the changing tropism of the virus ranging from primary cultures to a diverse continuum of animal models ^3,8,13,14^. Our current observations on the tropism change in primary swabs and low passage BA.5 isolates is also consistent with recent observation of increased replication in lung tissue and greater disease severity in animal models using BA.5 (**doi:** https://doi.org/10.1101/2022.05.26.493539).

To conclude, Omicron BA.5, like other omicron lineages, represents a continuing challenge for present vaccine strategies. Whilst many clinical studies have observed lower disease severity following infection in vaccine populations, the relative contributions of the Omicron variant phenotype and the efficacy of vaccine response to this observation was not readily clear ^14^. Whilst BA.1 and BA.2 are different in their transmissibility, their tropism towards cells of the upper respiratory tract is very similar. Mechanistically this change in tropism that moved efficient viral replication from the lung to bronchus is currently hypothesised to be a consequence of alternate protease usage away from the serine protease TMPRSS2 ^3,7,8,13^. Curiously, the efficient use of TMPRSS2 is associated with prior furin cleavage of the S1 and S2 domains of Spike ^3,12^. The furin cleavage site is retained in Omicron lineages and acts as a functional substrate for furin when expressed as a separate peptide from the Spike (https://doi.org/10.1101/2022.04.20.488969). Therefore, conformational changes within the Spike of BA.1 and BA.2 would be consistent with the furin cleavage site being held in a conformation with limited furin accessibility. In contrast, the small changes in the RBD of BA.5 appear to encourage furin cleavage in recent studies using pseudotyped viral platforms ^15^. This is consistent with observations for more efficient TMPRSS2 use, both herein and in animal models observing increased infection of the lung, in addition to increased disease severity ^16^. Curiously, the change in BA.5 not only gives the virus greater antibody evasion potential but concurrently has changed its tropism along with an increased transmission potential in the community. Whilst observations of disease severity initially in South Africa were similar to other Omicron lineages ^17^, the increasing prevalence of BA.5 globally will now enable observations of disease severity across a range of immune backgrounds. Unfortunately, as we have observed herein, prior Omicron infections in those post-three vaccine doses have primarily increased potent responses to early pre-Omicron variants and have done little to increase breadth across Omicron lineages. Moving forward, vaccine strategies that can rapidly come online to increase breadth to current variants or can induce responses to invariant epitopes that can future proof communities to emerging variants would be more pragmatic. In addition, whilst BA.5 appears to have evolved tropism towards pre-Omicron variant entry pathways, we will now need to look closely towards future variant trajectories and importantly if their tropism aligns with greater disease severity. If increased TMPRSS2 use is key to ranking variants for potential disease severity in the community, assays both herein and elsewhere will be able to resolve this changing TMPRSS2 tropism as soon as variants appear within the community through analysis of viral infectivity in primary nasopharyngeal swabs across TMPRSS2 cell lines.

## Materials & Methods

### Human sera and ethics statement

The ADAPT cohort is composed of RT-PCR–confirmed convalescent individuals (incl. some subsequently vaccinated) recruited in Australia since 2020 ^10^. The ADAPT cohort is composed of RT-PCR–confirmed convalescent individuals (incl. some subsequently vaccinated) recruited in Australia since 2020 ^10^. Serum from healthy volunteers vaccinated with ChAdOx1 and BNT162b2 was collected 4 weeks post second-dose vaccination. All human serum samples were obtained with written informed consent from the participants (2019/ETH03336; 2020/ETH00964; 2020/ETH02068; 2021/ETH00180).

### Other immunoglobulin products

Clinical grade Sotrovimab (62.5mg/ml; NDC 0173-0901-86) was kindly provided by GSK Healthcare while clinical grade Cilgavimab and Tixagevimab (100mg/ml each; AstraZeneca) were kindly provided by Dr Sarah Sasson (Kirby Institute, UNSW). Cilgavimab and Tixagevimab were mixed in equal volumes to generate the monoclonal antibody cocktail (Evusheld). All monoclonal antibodies were tested at a starting concentration of 10ug/ml and diluted two-fold in an eight step dilution series.

### Polyclonal Immunoglobulin preparations and anti-SARs-CoV-2 hyperimmune globulin

The immunoglobulins used herein were purified using the licensed and fully validated immunoglobulin manufacturing process used for Privigen (Stucki 2008), notionally similar to others (Karbiener et al. 2021). Thirteen Poly-Ig lots were manufactured using the Privigen process described by Stucki et al. (Stucki 2008) included US plasma collected by plasmapheresis from a mixture of vaccinated with SARS-CoV-2 mRNA vaccines, convalescent and non-convalescent donors (source plasma, n between 9495-23,667 per batch) collected in the September 2021. The WHO international reference standard for SARS-CoV-2 neutralization (NIBSC 20/136) was obtained from ^18^.

### Cell culture

HEK293T cells stably expressing human ACE2 and TMPRSS2 were generated by lentiviral transductions as previously described ^7 10^. A highly permissive clone (HAT-24) was identified through clonal selection and used for this study. The HAT-24 line has been extensively cross-validated with the VeroE6 line (manuscript submitted). HAT-24 cells and VeroE6-TMPRSS2 (CellBank Australia, JCRB1819) were cultured in DMEM-10%FBS and VeroE6 cells (ATCC® CRL-1586™) in MEM-10%FBS. All cells were incubated at 37°C, 5% CO_**2**_ and >90% relative humidity.

### Viral isolation, propagation, and titration

All laboratory work involving infectious SARS-CoV-2 occurred under biosafety level 3 (BSL-3) conditions. Diagnostic respiratory specimens testing positive for SARS-CoV-2 (RT-qPCR, Seegene Allplex SARS-CoV-2) were sterile-filtered through 0.22 μm column-filters at 10,000x g and serially diluted (1:3) on HAT-24 cells (10^**4**^ cells/well in 96-well plates). Upon confirmation of cytopathic effect by light microscopy, 300μL pooled culture supernatant from infected wells (passage 1) were added to VeroE6 cells in a 6-well plate (0.5 × 10^**6**^ cells/well in 2mL) and incubated for 48h. The supernatant was cleared by centrifugation (2000 xg for 5 minutes), frozen at -80 °C (passage 2), then thawed and titrated to determine median tissue culture infective dose (TCID50) on VeroE6-TMPRSS2 cells according to the Spearman-Karber method ^19^. Viral stocks used in this study correspond to passage 3 virus, which were generated by infecting VeroE6-TMPRSS2 cells at MOI=0.025 and incubating for 24h before collecting, clearing, and freezing the supernatant as above. Sequence identity and integrity were confirmed for both passage 1 and passage 3 virus via whole-genome viral sequencing using an amplicon-based Illumina sequencing approach, as previously described ^20^. For a list of the viral variants used in this study see Supplementary table S4. Passage 3 stocks were titrated by serial dilution (1:5) in DMEM-5%FBS, mixing with HAT-24 cells live-stained with 5% v/v nuclear dye (Invitrogen R37605) at 1.6×10^**4**^/well in 384-well plates, incubating for 20h, and determining whole-well nuclei counts with an InCell Analyzer high-content microscope and IN Carta analysis software (Cytiva, USA). Data was normalised to generate sigmoidal dose-response curves (average counts for mock-infected controls = 100%, and average counts for highest viral concentration = 0%) and median lethal dose (LD_**50**_) values were obtained with GraphPad Prism software.

### Rapid high-content SARS-CoV-2 microneutralization assay with HAT-24 cells (R20)

Human sera or monoclonal antibodies were serially diluted (1:2 series starting at 1:10) in DMEM-5%FBS and mixed in duplicate with an equal volume of SARS-CoV-2 virus solution standardised at 2xLD_**50**_. After 1h of virus–serum coincubation at 37°C, 40μL were added to an equal volume of nuclear-stained HAT-24 cells pre-plated in 384-well plates as above. Plates were incubated for 20h before enumerating nuclear counts with a high-content fluorescence microscopy system as indicated above. The % neutralization was calculated with the formula: %N = (D-(1-Q)) × 100/D as previously described ^10^. Briefly, “Q” is a well’s nuclei count divided by the average count for uninfected controls (defined as having 100% neutralization) and D = 1-Q for the average count of positive infection controls (defined as having 0% neutralization). Sigmoidal dose-response curves and ID_**50**_ values (reciprocal dilution at which 50% neutralization is achieved) were obtained with GraphPad Prism software. Neutralization assays with VeroE6 cells were performed exactly as described above excepting that; input virus solution was standardised at 1.25 × 10^**4**^ TCID50/mL, cells were seeded at 5×10^**3**^ cells/well in MEM-2%FBS (final MOI = 0.05), plates were incubated for 72h, and cells were stained with nuclear dye only 1h before imaging.

### End Point titers for primary Nasopharyngeal Swabs

Nasopharyngeal swabs were collected in Edwards virus transport media (Narrellan, NSW, Australia) for the purpose diagnostic testing at Molecular Diagnostic Medicine Laboratory, Sydpath, St Vincent’s Hospital, Sydney. The use of residual diagnostics samples to isolated and/or study emerging variants was independently reviewed and approved by the New South Wales Chief Health Officer. 300ul of SARS CoV-2 PCR positive samples were then aliquoted and frozen at -80 for latter use. As outlined above, samples were 0.22 μm column-filtered at 10,000x g for 10’ at 4°C. Samples were then added to 5000 HAT-24 cells in 80ul total across a 384 well plate. Each sample and dilutions were run in quadruplicate and scored for cytopathic effect by light microscopy. End point titers were then calculated using Spearman-Karber method ^19^. For direct comparison of BA2 and BA5 within the community, samples were collected over the month of June 2022, when BA5 was starting to supplant BA2. As there were primarily two viral isolates circulating, we resolved viral genomes through nanopore sequencing as previously described ^20^ concurrently when determining viral end point point titers. Samples of pre-Omicron variants are described elsewhere ^7^, were derived from primary samples where 95% were from unvaccinated patients and are presented as a comparison for Omicron lineages BA1, BA2 and BA5.

### Immunophenotyping of ACE2 and TMPRSS2

HEK293T, HAT-24 and Calu-3 cells were assessed by flow cytometry using either (i) unconjugated rabbit anti-TMPRSS2 antibodies (Novus) followed by secondary goat anti-rabbit AlexaFluor594 (ThermoFisher) or (ii) directly conjugated ACE2-AlexaFluor647 antibodies (R&D Systems). Cells were acquired with a BD Fortessa flow cytometer and analysed using FlowJo software (v 10.8.0, BD Biosciences).

## Data Availability

All data produced in the present study are available upon reasonable request to the authors.
Upon publication we will provide data through Figshare

## Supplementary Tables

Excel spreadsheet showing supplementary Tables 1-3

Supplementary Table S4. SARS-CoV-2 variants used in this study.

| Variant | Lineage | Seq ID | GISAID ID |
| --- | --- | --- | --- |
| Ancestral | A.2.2 | nCoV_2715 | hCoV-19/Australia/NSW4715/2021 |
| Beta | B.1.351 | nCoV_2463 | hCoV-19/Australia/NSW4463/2021 |
| Delta | B.1.617.2 | nCoV_2605 | hCoV-19/Australia/NSW4605/2021 |
| Omicron BA.1 | B.1.1.529 | RCoV_1933 | hCoV-19/Australia/NSW-RPAH-1933/2021 |
| Omicron BA.2 |  | Pending | Pending |
| Omicron BA.5 |  | Pending | Pending |

## References Cited

1 Rajah, M. M. et al. SARS-CoV-2 Alpha, Beta, and Delta variants display enhanced Spike-mediated syncytia formation. EMBO J 40, e108944, doi:10.15252/embj.2021108944 (2021).

2 Zhao, H. et al. SARS-CoV-2 Omicron variant shows less efficient replication and fusion activity when compared with Delta variant in TMPRSS2-expressed cells. Emerg Microbes Infect 11, 277–283, doi:10.1080/22221751.2021.2023329 (2022).

3 Meng, B. et al. Altered TMPRSS2 usage by SARS-CoV-2 Omicron impacts infectivity and fusogenicity. Nature 603, 706–714, doi:10.1038/s41586-022-04474-x (2022).

4 Halfmann, P. J. et al. SARS-CoV-2 Omicron virus causes attenuated disease in mice and hamsters. Nature 603, 687–692, doi:10.1038/s41586-022-04441-6 (2022).

5 Diamond, M. et al. The SARS-CoV-2 B.1.1.529 Omicron virus causes attenuated infection and disease in mice and hamsters. Res Sq, doi:10.21203/rs.3.rs-1211792/v1 (2021).

6 Suzuki, R. et al. Attenuated fusogenicity and pathogenicity of SARS-CoV-2 Omicron variant. Nature 603, 700–705, doi:10.1038/s41586-022-04462-1 (2022).

7 Aggarwal, A. et al. Platform for isolation and characterization of SARS-CoV-2 variants enables rapid characterization of Omicron in Australia. Nat Microbiol 7, 896–908, doi:10.1038/s41564-022-01135-7 (2022).

8 Hui, K. P. Y. et al. SARS-CoV-2 Omicron variant replication in human bronchus and lung ex vivo. Nature 603, 715–720, doi:10.1038/s41586-022-04479-6 (2022).

9 Dejnirattisai, W. et al. SARS-CoV-2 Omicron-B.1.1.529 leads to widespread escape from neutralizing antibody responses. Cell 185, 467–484 e415, doi:10.1016/j.cell.2021.12.046 (2022).

10 Tea, F. et al. SARS-CoV-2 neutralizing antibodies: Longevity, breadth, and evasion by emerging viral variants. PLoS Med 18, e1003656, doi:10.1371/journal.pmed.1003656 (2021).

11 Pinto, D. et al. Cross-neutralization of SARS-CoV-2 by a human monoclonal SARS-CoV antibody. Nature 583, 290–295, doi:10.1038/s41586-020-2349-y (2020).

12 Mlcochova, P. et al. SARS-CoV-2 B.1.617.2 Delta variant replication and immune evasion. Nature 599, 114–119, doi:10.1038/s41586-021-03944-y (2021).

13 Shuai, H. et al. Attenuated replication and pathogenicity of SARS-CoV-2 B.1.1.529 Omicron. Nature 603, 693–699, doi:10.1038/s41586-022-04442-5 (2022).

14 Kozlov, M. Omicron’s feeble attack on the lungs could make it less dangerous. Nature 601, 177, doi:10.1038/d41586-022-00007-8 (2022).

15 Zhang, Y. et al. SARS-CoV-2 spike L452R mutation increases Omicron variant fusogenicity and infectivity as well as host glycolysis. Signal Transduct Target Ther 7, 76, doi:10.1038/s41392-022-00941-z (2022).

16 Balint, G., Voros-Horvath, B. & Szechenyi, A. Omicron: increased transmissibility and decreased pathogenicity. Signal Transduct Target Ther 7, 151, doi:10.1038/s41392-022-01009-8 (2022).

17 Tegally, H. et al. Emergence of SARS-CoV-2 Omicron lineages BA.4 and BA.5 in South Africa. Nat Med, doi:10.1038/s41591-022-01911-2 (2022).

18 Kristiansen, P. A. et al. WHO International Standard for anti-SARS-CoV-2 immunoglobulin. Lancet 397, 1347–1348, doi:10.1016/S0140-6736(21)00527-4 (2021).

19 Ramakrishnan, M. A. Determination of 50% endpoint titer using a simple formula. World J Virol 5, 85–86, doi:10.5501/wjv.v5.i2.85 (2016).

20 Bull, R. A. et al. Analytical validity of nanopore sequencing for rapid SARS-CoV-2 genome analysis. Nat Commun 11, 6272, doi:10.1038/s41467-020-20075-6 (2020).

