## Supplementary Figure for "SARS-CoV-2 Omicron BA.5: Evolving tropism and evasion of potent humoral responses and resistance to clinical immunotherapeutics relative to viral variants of concern"

Supplementary figure 1: Expression of ACE2 and TMPRSS2 in Hek 293T, HAT-24 and Calu-3

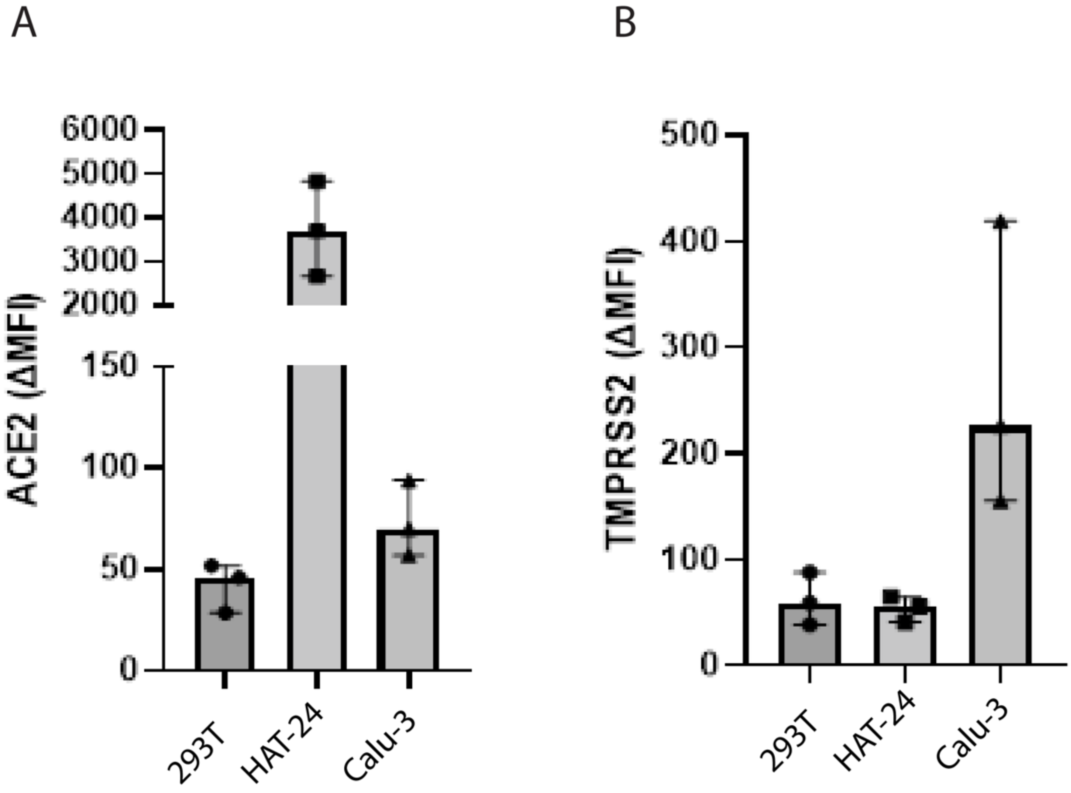


Figure S1: Expression ACE2 (A) and TMPRSS (B) was examined in Hek293T, ACE2-TMPRSS2 293T (HAT-24) and Calu-3 cells was examined using flow cytometry. Shown are the mean and standard deviations from three independent experiments.
